# Quality assessment of studies included in Cochrane oral health systematic reviews

**DOI:** 10.1101/2020.10.10.20210518

**Authors:** Ahmad Sofi-Mahmudi, Pouria Iranparvar, Maryam Shakiba, Erfan Shamsoddin, Hossein Mohammad-Rahimi, Sadaf Naseri, Parisa Motie, Bita Mesgarpour

**Author notes:** Corresponding author: Pouria Iranparvar; Address: 21, National Institute for Medical Research Development (NIMAD), West Fatemi St., Tehran, Tehran, Iran; Postal code: 1419693111. The authors of this study explicitly claim that there was no conflict of interest in conducting this work. Funding source: This research did not receive any specific grant from funding agencies in the public, commercial, or not-for-profit sectors. **Author contributions:** AS-M conceived, designed, conducted, and supervised the study, interpreted the data, and wrote and revised the manuscript. PI conducted the study, interpreted the data, and wrote and revised the manuscript. MS conducted the study, interpreted the data, and wrote and revised the manuscript. ES wrote and revised the manuscript and interpreted the data. HM-R conducted the study, and revised the manuscript. SN conducted the study, and revised the manuscript. PM conducted the study, and revised the manuscript. BM wrote and revised the manuscript.

## Abstract

**Objectives:** The Risk of Bias (RoB) and other characteristics of randomized clinical trials included in Cochrane oral health systematic reviews were assessed.

**Study Design and Settings:** All the trials included in Cochrane oral health systematic reviews were examined. The RoB was evaluated for all the included clinical trials according to the Cochrane review standards. The Overall Risk of Bias (ORoB) was defined in this study based on the criteria for determining the overall bias in Cochrane’s RoB tool-v2. Descriptive analyses were carried out to determine the frequency of each intended variable.

**Results:** A total of 2565 studies were included in our analysis. The majority of the studies (n=1600) had sample sizes of 50 or higher. As for blinding, 907 studies were labelled as double-blind. Performance bias showed the highest rate of high risk (31.4%). Almost half of the studies had a high ORoB compared to 11.1% with low ORoB. The studies that used placebos had higher low ORoB (14.8% vs. 10.7%). The double-blind studies had the highest low ORoB (23.6%). The studies with a cross-over design had the highest low ORoB (28.8%).

**Conclusion:** Overall, the RoB for the studies on dentistry and oral health in Cochrane reviews was deemed high.

## Introduction

The quality assessment of studies should always consider both internal and external validities (1), which are critical aspects of any scientific project, although internal validity is more relevant to empirical studies (2). The Risk of Bias (RoB) is a good measure of the internal validity of a study (3, 4). Bias in clinical studies leads to the over- or under-estimation of treatment outcomes (5). From a clinical perspective, bias can lead to the application of an intervention that may not be effective or that may even be potentially harmful (3). While Randomized Clinical Trials (RCTs) are widely considered the gold standard for therapeutic clinical research to measure the effectiveness of new medical interventions (6), they are also prone to bias (6).

The Cochrane Database of Systematic Reviews is proposed as the biggest and most recognized scientific database of systematic reviews and meta-analyses in health sciences that publishes high-quality systematic reviews with specific methodological protocols (7). In the case of RoB, Cochrane reviews use a specific domain-based evaluation tool that has been changed over the past years, with the latest edition introduced in 2008 (8). Currently, the domains consist of ‘sequence generation’, ‘allocation concealment’, ‘blinding of participants and personnel’, ‘other potential threats to validity (other sources of bias)’, ‘blinding of outcome assessment (blinding of the examiner)’, ‘incomplete outcome data’, and ‘selective outcome reporting’. Cochrane’s evaluation tool for RoB is a signposting scale with three variations, namely, ‘Low-risk’, ‘High-risk’ and ‘Unclear-risk’. The first two variations are self-explanatory, and the unclear-risk choice is assigned when there is not enough evidence available (reported by the study) to explicitly draw a conclusion about any specific domain. The primary sources of support and reliance for Cochrane judgments are pieces of evidence or quotes from the appraised paper or any rational inferences and reviews drawn from the author’s explanations (9).

Despite all the invaluable efforts made in the past few years, there is still a considerable gap between the available evidence and the common clinical procedures in dentistry (10). As a result, in many cases, it may be difficult for practitioners to extract their needed data from the relevant studies (11). Moreover, there are also studies with inadequate allocation concealment and high RoB, which tend to change empirical results in a clinical setting (12). Without a way to offer dentists high-quality evidence to reinforce their decision-making process, dentists will be faced with substantial stress and their decision-making will become more time-consuming in clinical settings. The present study aimed to assess the RoB and other characteristics of the RCTs included in Cochrane oral health systematic reviews on the subject of dentistry and oral health. Further suggestions to be considered by future Cochrane review teams will be presented toward the end.

## Methods

This study is a secondary analysis of the data provided in Cochrane oral health systematic reviews. All the judgements concerning the quality of the RCTs have been made by Cochrane review authors.

### Search strategy

The “dentistry and oral health” filter was used to search Cochrane Library’s “Search Reviews (CDSR)” section on June 1, 2020. All the reviews in this category were included in the data extraction step.

### Identification and selection of primary studies

The primary studies were identified using the references included in the systematic reviews and their data were extracted by the characteristics of the included studies’ tables. Figure 1 shows the detailed flow diagram of steps taken to choose the eligible studies.

**Figure 1.**
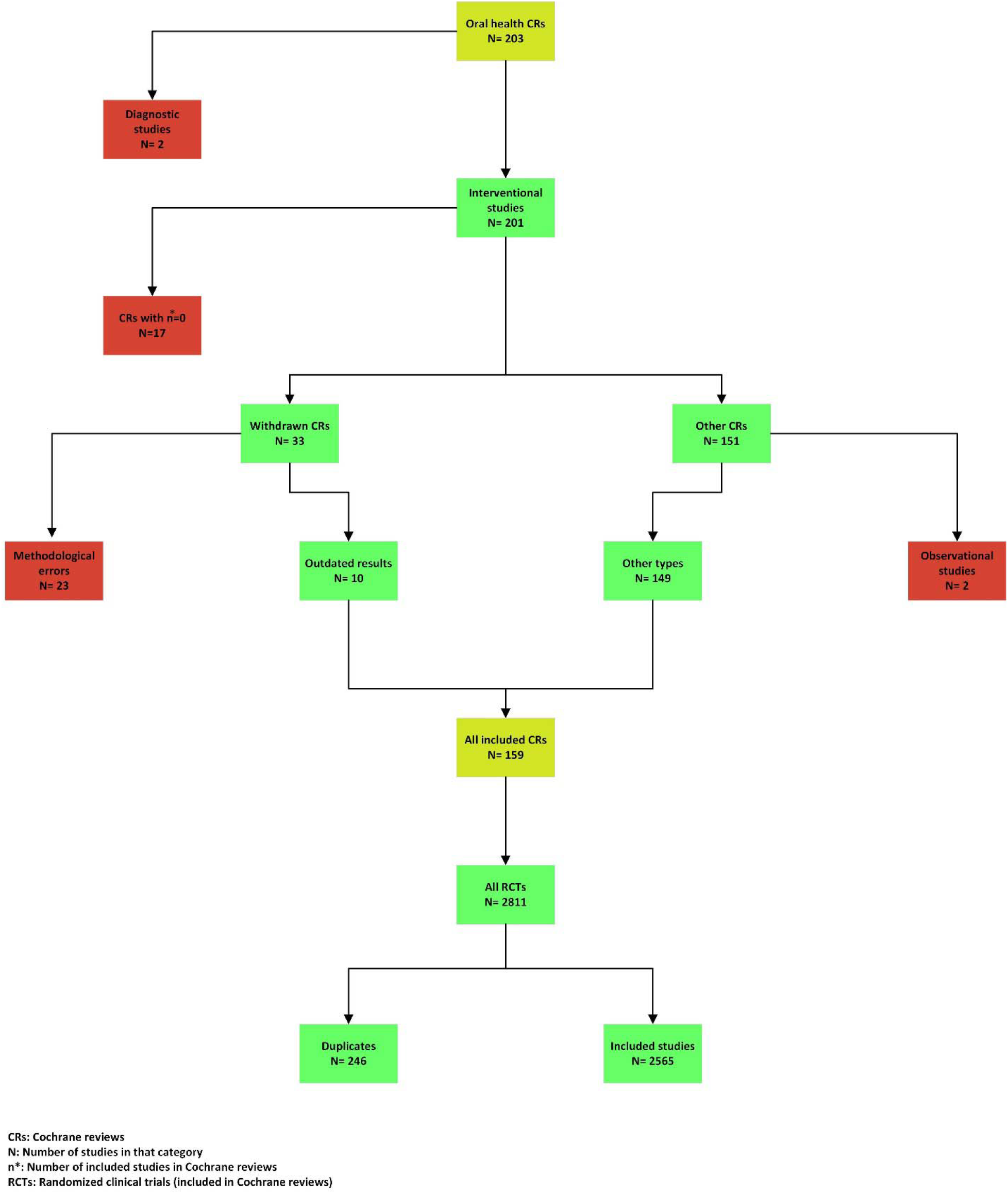
Flowchart for study selection strategy.

### Data extraction and management

This phase of the study was carried out for all the included trials by seven independent authors (AS-M, PI, MS, ES, HM, SN, PM), and the results were entered directly into preformatted Excel 2016 spreadsheets. All the spreadsheets were then merged into one file and rechecked to avoid any mistakes and inconsistencies among the data extractors (AS-M, PI, MS). The items intended to be extracted were divided into three categories: (1) Basic information on the RCTs, e.g. title, first author, year of publication, etc.; (2) The information reported in the RCTs (the “Characteristics of the included studies” section), which basically characterized the main methodological features of each RCT; and 3) The RoB assessment of the RCTs.

Given that the blinding process was evaluated differently by the Cochrane review authors, the RoB assessments were performed by different trajectories. In some studies, blinding took place in two subgroups, i.e. blinding of the participants and personnel (performance bias) and blinding of the outcome assessment (detection bias); meanwhile, in some other studies, blinding was taken as one item. In rare instances, blinding took place in two categories, including subjective and objective outcomes or a range, namely, assessor, analyst, participants, and caregivers. In these cases, an aggregated form was provided; if one or more subcategories were reported as high-risk, the whole category of blinding bias was considered high-risk, and if all were at a low risk, the whole category was considered low-risk; otherwise, the blinding bias was reported as unclear.

There were some other RoB assessment items that were rarely reported; for instance, funding bias, intention to treat bias, sample size bias, for-profit bias, and power calculation bias. Due to the low frequency of these items, these kinds of biases were not extracted in this study. Additionally, a new variable was defined to aggregate the full results of the assessments, named the “Overall Risk of Bias” (ORoB). The criteria for determining the ORoB were the same as those of the “Overall Bias” assessment in Cochrane’s RoB tool -version 2 (13). If any of the bias domains were considered high-risk, the ORoB was considered high. In the absence of high-risk biases, unclear-risk-assessed biases were perceived in the same way; otherwise, the ORoB was considered low. The RCTs that appeared in more than one Cochrane review were excluded if their RoB assessments were not consistent.

In including RCTs in this study, several groups were considered, namely, quasi-experimental (controlled before-after, interrupted time series, and non-randomized designs), parallel RCT, crossover RCT, cluster RCT, split-mouth RCT, and repeated-measures study designs.

Finally, to scrutinise the Cochrane review authors’ subjectiveness and bias of judgement, the studies were distinguished by RoB domains. Cochrane reviews that included the same studies, either with the same or with different RoB assessments, were listed and signposted in different categories based on the included titles.

### Statistical analysis

The extracted data were analysed using R v3.6.0 (2019-04-26) (R Core Team, R Foundation for Statistical Computing, Vienna, Austria. http://www.r-project.org), and mean and standard deviations were reported for the quantitative continuous variables and frequencies for the qualitative data.

## Results

### Characteristics of Cochrane Reviews

Of the 8293 Cochrane reviews on the subject, 203 (2.4%) were on dentistry and oral health (23^rd^ rank among the subjects in terms of the number of reviews). The majority of the reviews were interventional (99.0%) and there were two diagnostic studies. The review subjects were mainly concerned with dental caries (24.6%), craniofacial anomalies (18.7%), and oral and maxillofacial surgery (15.8%). Seventeen reviews (8.4%) included zero studies and 33 (16.3%) reviews were withdrawn for different reasons, mainly because of being out-of-date (75.8%) and not meeting the current Cochrane methodological standards (54.5%). Nonetheless, ten of the withdrawn studies were included as they were still methodologically sound. Besides, two reviews included only observational studies. Hence, 159 interventional reviews (80.3%) provided the data needed for this study.

The mean and median of the number of the studies included in these reviews were 18.7 (SD=26.0) and 10 (range: 1-154), respectively. Four reviews (4/203, 2.0%) had more than 100 included studies and 80 (80/203, 39.4%) had less than ten (excluding the zero-including studies). The subjects of three most-cited reviews were oral cancer and pre-cancerous lesions (N=309), oral and maxillofacial surgery (N=305), and dental caries (N=275) according to Web of Science. Nonetheless, reviews about antibiotic therapy (78.5, N=2), oral lichen planus (73.5, N=2), and periodontal diseases (69.6, N=17) had the highest mean number of citations.

### Characteristics of the included studies

The reviews included 2811 studies in which 246 were duplicated. Therefore, a total of 2565 studies were included in our analysis. Almost two-thirds of these studies (n=1666) were published after 2000. The distribution of the location where the studies were conducted was quite heterogeneous. The USA, the UK, and India had the highest number of studies with 588 (22.9%), 221 (8.6%), and 164 (6.4%) studies, respectively. Sixty-eight studies were conducted using international collaborations. Figure 2 shows the distribution of the location of the studies.

**Figure 2.**
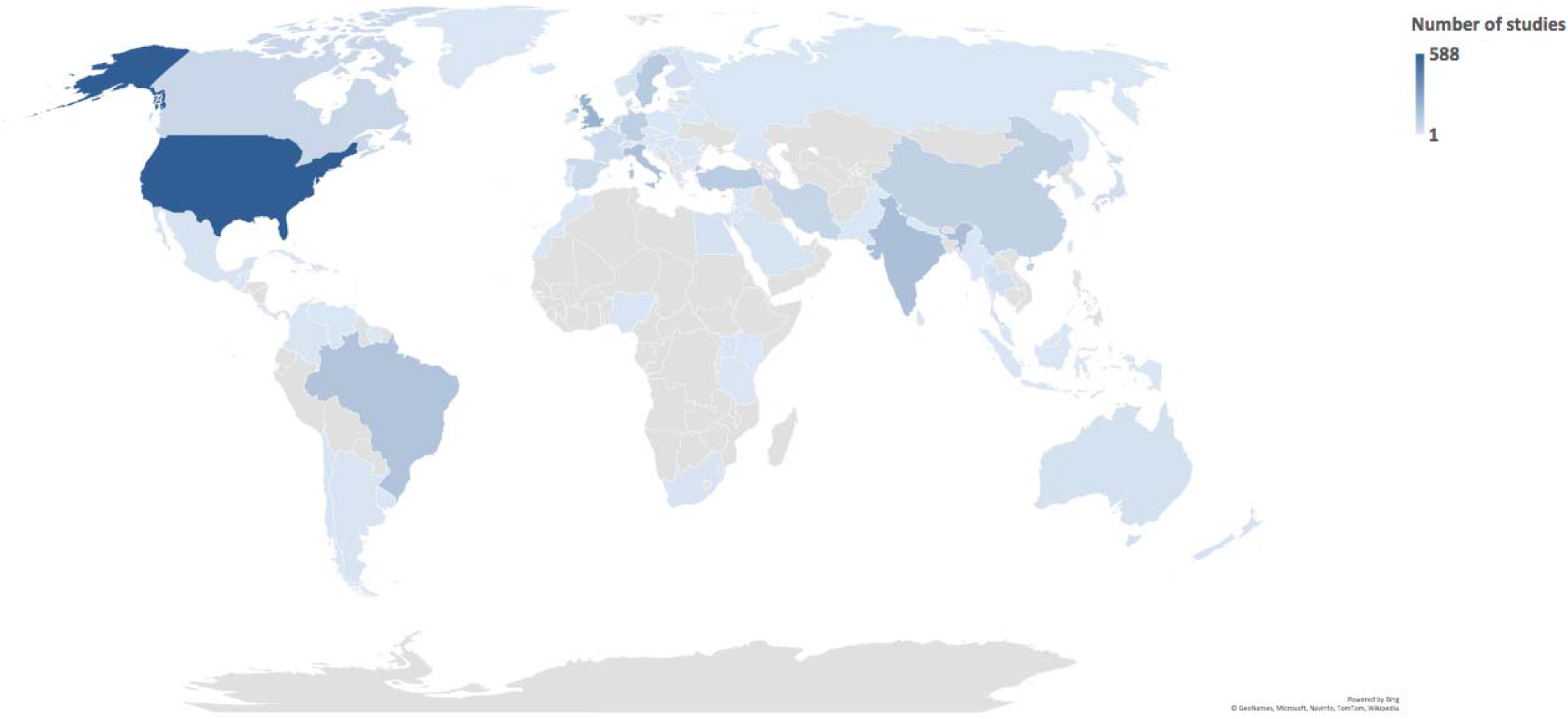
World map for the distribution of conducting sites.

Out of the 614 study sources, four were unpublished and one was a dissertation from the University of Sao Paulo, Brazil. However, 117 journals had more than three studies included in the Cochrane reviews. Most of these journals were published in the USA (47.9%) and the UK (26.4%). According to the SCImago Journal Rank (SJR) quartiles (2019), over half of these journals were categorized as Q1 (52.1%), followed by Q2 (25.6%). The most frequent subjects of the journals were dentistry (miscellaneous; 41.0%), followed by medicine (miscellaneous; 10.3%) and oral surgery (10.3%). The following journals represented the highest inclusions in the Cochrane reviews: Journal of Clinical Periodontology (n=108, 4.2%), and Journal of Periodontology (n=103, 4.0%), and Journal of the American Dental Association (n=67, 2.6%).

The majority of the studies (n=1600, 62.4%) had sample sizes of 50 or higher. The mean and median of the sample sizes were 1017 (SD=3875.1) and 303 (range: 3-191873), respectively, and 86 studies had sample sizes higher than 1000. In contrast, 4.3% of the studies had less than 20 participants. The sample size was not reported in the tables in 17 studies.

More than three-fourths of the included studies were parallel RCTs (n=2040), followed by split-mouth (n=253), cross-over (n=154), and quasi-experimental (n=72) studies. The design was unclear in one study.

From a methodological point of view, 787 (30.7%) studies reported that they had one arm as the control group that received placebo. As for blinding, 907 (35.3%) studies were labelled as double-blinded, 60 studies (2.4%) as triple-blinded, and 425 (16.6%) studies had not used any blinding in their design. The number of double- and triple-blinded studies was 967, and 791 of them (81.8%) had low risks in terms of performance bias.

Mostly all the domains of the RoB tool were assessed in all the reviews. However, in 45 (1.8%) and 11 (0.4%) of the interventional studies, respectively, random sequence generation and allocation concealment were not assessed by the reviewers. Among the various domains of bias, performance bias (blinding of the participants and personnel) showed the highest rate of high risk (31.4%). Instead, other sources of bias had the highest rate of low risk (68.2%). Figure 3 presents the overall assessment of each domain of the RoB tool. According to Figure 4, random sequence generation data has continuously improved in terms of increasing low RoB during 2000-2019.

**Figure 3.**
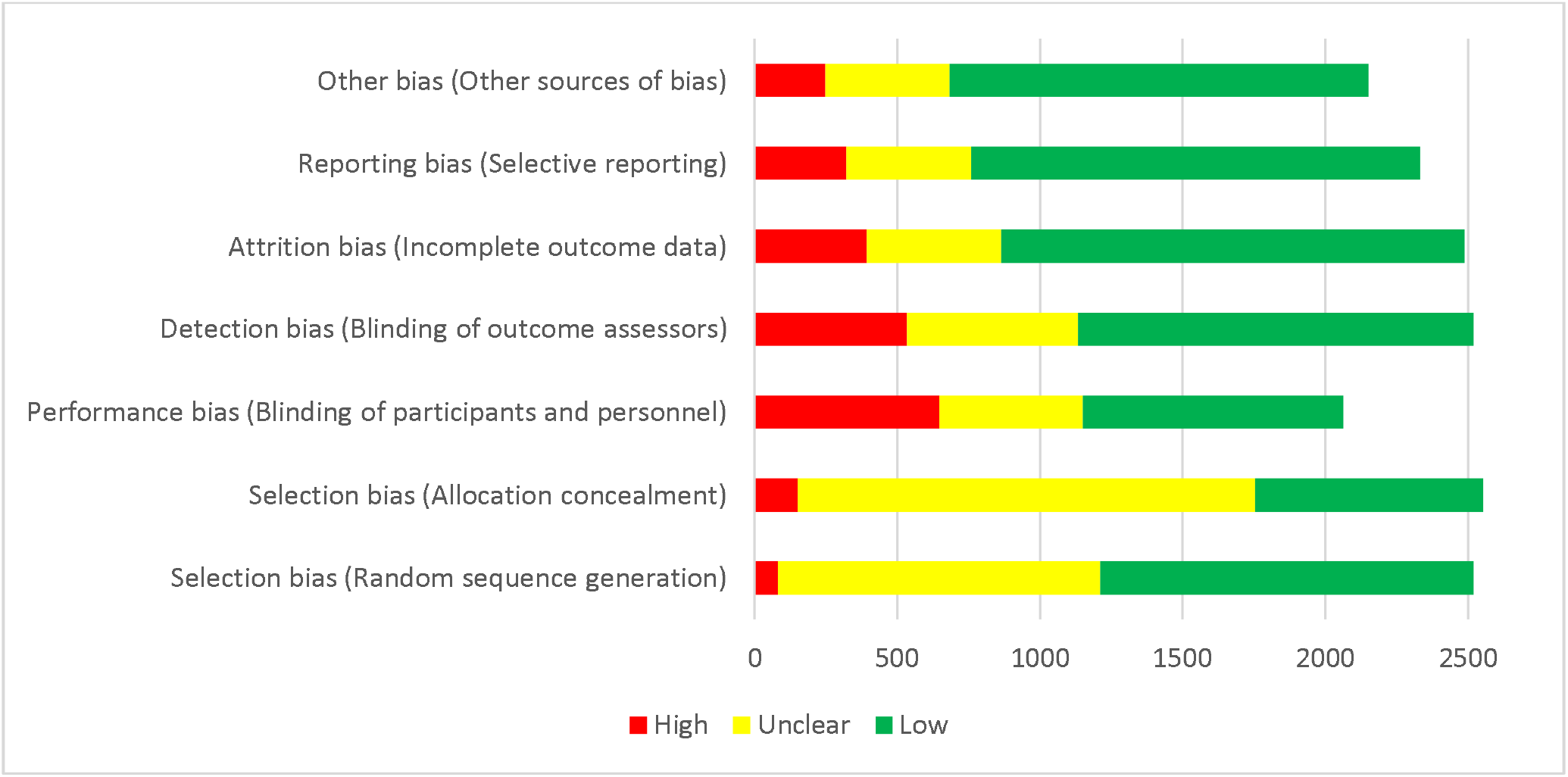
Risk of Bias assessment for all domains of RCTs included and evaluated in Cochrane Oral Health systematic reviews

**Figure 4.**
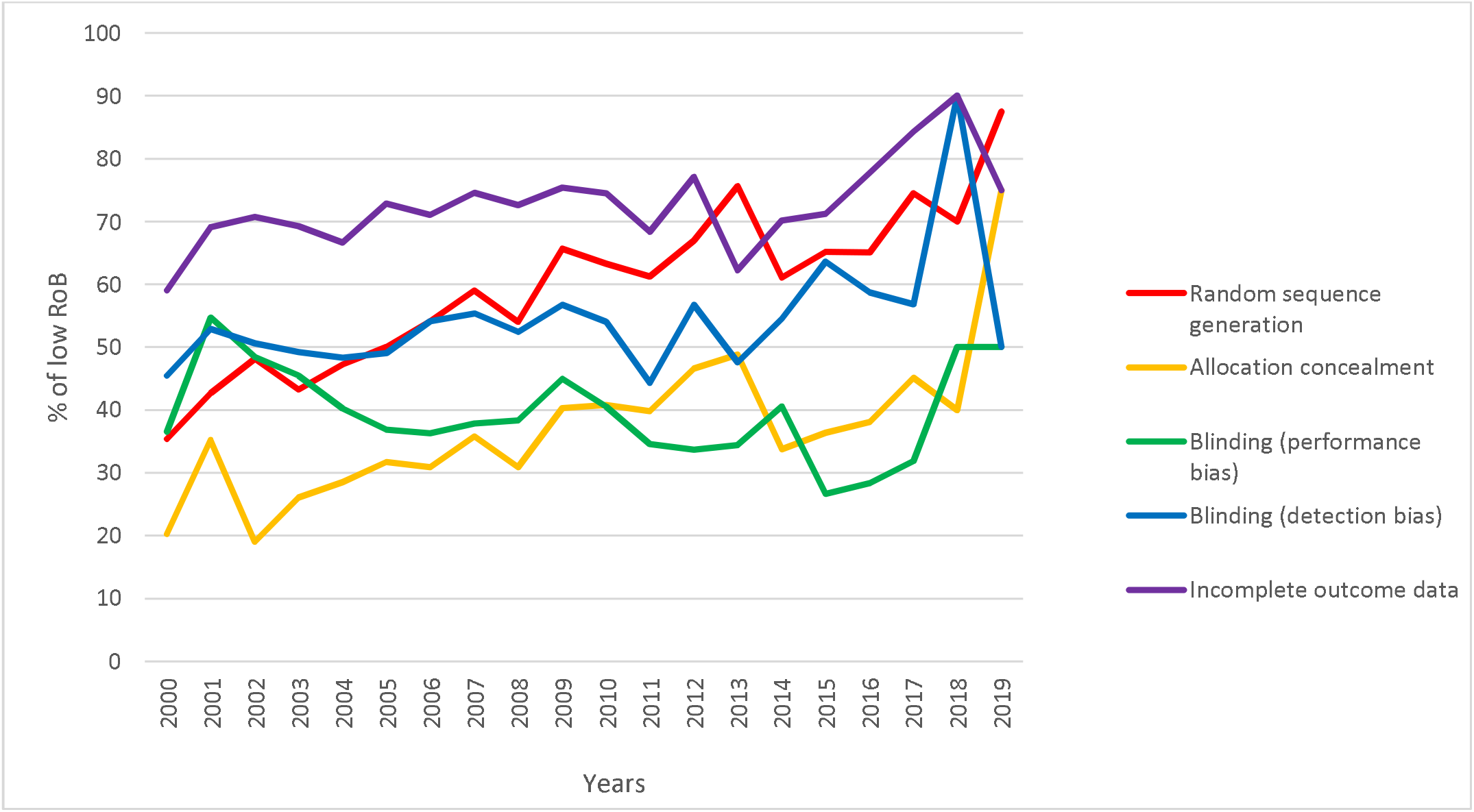
The trend of low RoB for five domains of Cochrane tool in oral health RCTs during 2000-2019

One hundred and forty-four studies had appeared in more than one Cochrane review (out of a total of 390 studies), with inconsistent RoB assessments in 107 (74.3%) of them. The assessment of attrition bias (incomplete outcome data; n=207, 53.1%) and selection bias (allocation concealment; n=34, 8.7%) had the highest and lowest rates of disagreement between the different reviews, respectively.

### The Overall Risk of Bias (ORoB) and its association with the study characteristics

After removing the duplicate studies that had not reached consistent assessments concerning the RoBs (determined in the Cochrane reviews), 2458 studies remained for calculating the ORoB. Near half of these studies (n=1119) had a high ORoB compared to 11.9% (n=294) with a low ORoB.

The following journals showed the highest percentage of studies with low ORoBs (considered eligible with at least ten included studies): The European Journal of Oral Implantology (73.1% of 26 studies), Journal of Endodontics (52.3%, of 44 studies), and Anesthesia Progress (38.5%, of 13 studies). The following journals showed the highest percentage of studies with high ORoBs (considered eligible with at least ten included studies): Journal of Dentistry for Children (91.7% of 12 studies), European Archives of Paediatric Dentistry (81.8%, of 11 studies), and Cancer (74.2%, of 31 studies).

Among the countries with more than 50 included studies, Italy and China had the highest percentage of low (32 out of 155, 20.6%) and high (49 out of 89, 55.1%) ORoBs, respectively.

Of the 821 studies conducted before the year 2000, 6.6% had low ORoBs, compared to 14.5% of those conducted after 2000 and 15.5% of those conducted after 2010. The studies that used placebos had higher percentages of low ORoB (14.8% vs. 10.7%) and lower percentages of high ORoB (27.6% vs. 52.7%). The double- and triple-blind studies had higher percentages of low ORoB (23.6% and 23.3%, respectively) while the non-blinded studies had the lowest percentage of low ORoB (1.0%) and highest percentage of high ORoB (80.4%). The studies with a cross-over design had the highest percentage of low ORoB (28.8%), followed by cluster RCTs (11.9%) and parallel studies (11.2%). The quasi-experimental mainly had a high ORoB (98.4%).

The high-ORoB studies had the highest mean sample size (n=414.3), followed by the unclear (n=213.6) and low (n=190.0) ORoBs.

## Discussion

Ranked as 23^rd^ among the 37 subjects in terms of volume, the “dentistry and oral health” subject comprises 2.4% of all Cochrane reviews. There is thus still a clear paucity of dental evidence in both interventional and diagnostic aspects compared to other medical disciplines(14). Aside from the quantity, the quality of the research is paramount and needs to be continuously enhanced as well. The dearth of evidence in dentistry and oral health subjects intensifies the need for high-quality studies on which clinicians can rely. To evaluate the quality of dental clinical trials, the RoB and other characteristics of the studies included in the Cochrane oral health systematic reviews were evaluated.

Approximately half of the clinical trials (47.3%) showed a high ORoB compared to 11.1% with a low ORoB. This finding was in line with the results reported by Yordanov et al., which revealed 43% of high ORoBs in medical clinical trials (15). Earlier assessments of RoB in dental clinical trials showed a significant decrease in studies judged as having inadequate methodological standards using the Cochrane risk of bias tool (16). Meanwhile, no specific associations were identified to guide dental practitioners for optimizing their future trials. The present findings are highly suggestive of using control groups in oral health-related interventional studies as it could noticeably decrease the ORoB. Furthermore, double- and triple-blind oral health-related interventional studies showed lower ORoBs compared to unblinded studies. Quasi-experimental studies showed higher ORoBs compared to cross-over, cluster RCT, and parallel designs. These findings can ultimately assist clinical researchers to direct their studies in more organized settings and designs and consider minimizing probable RoBs in their relevant projects.

Even though dentists are able to conduct high-quality studies with low risks of performance bias (95.9% of the double- and triple-blinded studies), apparently they have not been very eager to follow the blinding protocols of clinical interventional studies (cumulative percentage of double- and triple-blinded studies=38.5%). This finding might be an implicit sign of incomprehensive dental research teams and the exclusive dispositions of clinical research in dentistry. Changing the exclusivity trend in research teams is proved to mainly impact the quality of evidence (17, 18).

Despite knowing that using control groups decreases the ORoB, only 32% of the clinical trials reported an arm as a control group. Also, among the various RoB domains, performance bias (blinding of the participants and personnel) showed the highest rate of high risk (31.4% of the studies). Consequently, setting correct methodological strategies and outlining the reporting protocol before conducting the study are imperative as well (15). These goals can be ensured by seeking the professional help of clinical methodologists and consulting with statisticians when planning studies and outlining their stages (17). Providing better platforms for these communications and constantly promoting their feasibility features are critical (19). Another long-term solution for decreasing the RoBs is to enhance the knowledge of dental students and dental practitioners about research methods (14, 20, 21). Teaching research methods should be pursued in parallel with the provision of more feasible and convenient ways for dental clinicians to contact professional methodologists, and a recommended step is to consult a statistician in every medical study since they can help refine the study design before the practical steps are taken. The patients are the ultimate stakeholders of these evidence-based decision-makings by dental practitioners, since they can be provided with the most reliable and up-to-date therapeutic options (22-24).

This study takes into account the fact that the blinding of the care providers is unachievable in some clinical trials in dentistry (e.g. surgical cases). Nonetheless, there are still various aspects of dental clinical trials that could potentially be improved by simple feasible actions, such as the accurate reporting of sequence generation and allocation strategies or using the most relevant yet feasible study designs.

## Conclusion

Nearly half of the dentistry and oral health-related clinical trials (47.3%) showed high ORoBs. Consequently, the quality of this dental evidence needs further enhancement and is inadequate in a noticeable part. If applicable, using methodological quality standards (e.g. blinding of the participants, using control groups, etc.) and using pre-determined study protocols (before conducting the research) and refining study designs to the most randomized settings possible can decrease the ORoB. Providing more feasible communication platforms for use among clinical researchers and statisticians and enhancing research methodology knowledge among clinicians can ultimately pave the way for conducting more high-standard studies in dentistry.

**Table 1.**
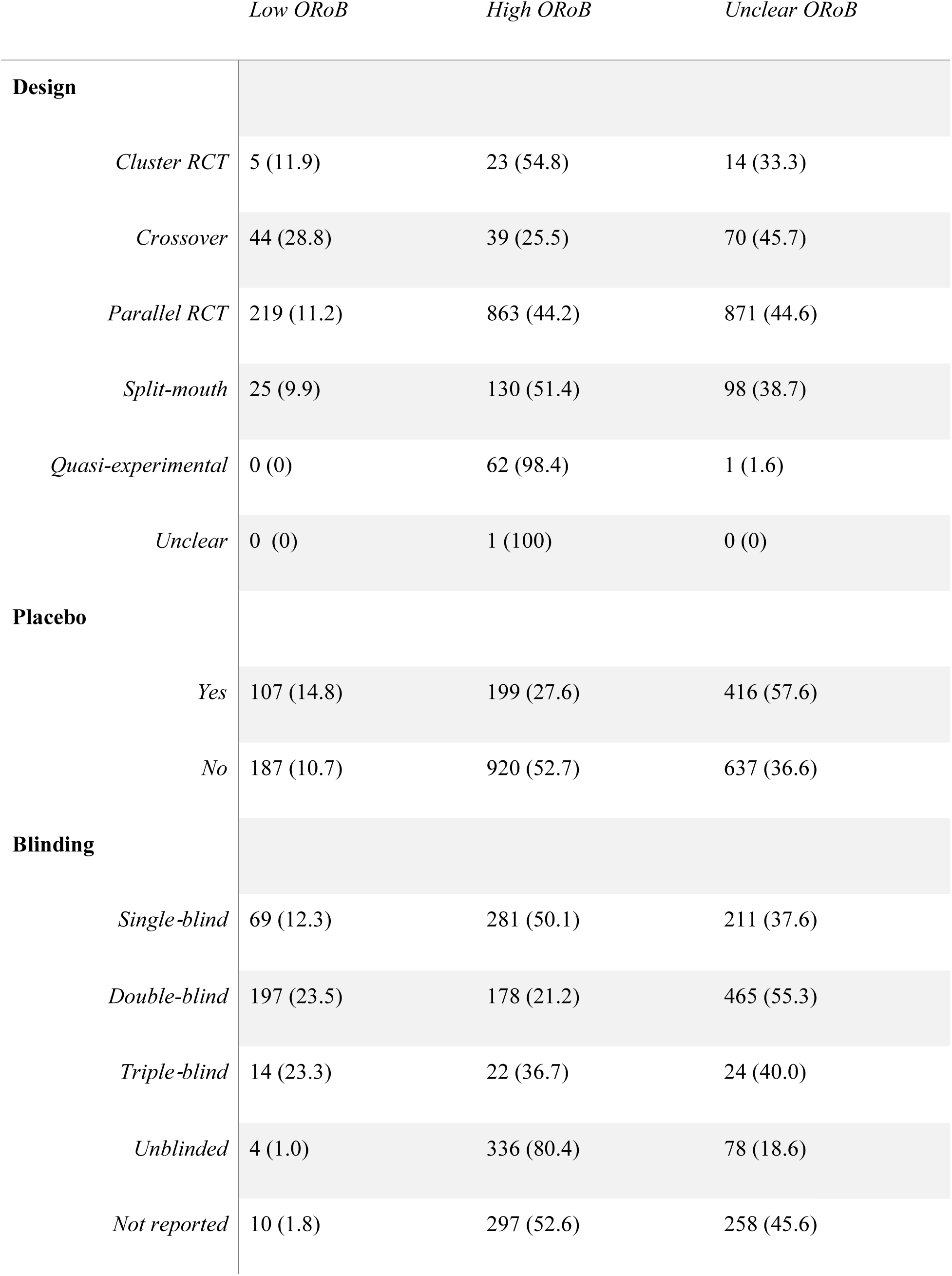
ORoB for each study design and method.

## Data Availability

Data is available at: https://doi.org/10.6084/m9.figshare.13070135.v2

## Availability of data and materials

The datasheet is available at https://doi.org/10.6084/m9.figshare.13070135.v2

